# Modeling the impact of screening, vaccination and treatment on the transmission dynamics of HPV and Cervical cancer

**DOI:** 10.64898/2025.12.09.25341930

**Authors:** Grace Mbugua, Caroline Kanyiri

**Author notes:** These authors contributed equally to this work.

## Abstract

Cervical cancer remains a significant cause of mortality and economic burden, particularly in developing countries with low rates of human papillomavirus (HPV) vaccination and screening. To address this, we present a mathematical model for controlling cervical cancer by integrating strategic HPV vaccination, screening and treatment. The population is divided into seven compartment: susceptible, vaccinated, infected with HPV, screened, cervical cancer, under treatment, and recovered. The model’s well-posedness is first established by proving the boundedness and non-negativity of solutions, ensuring biological relevance. The basic reproduction number *R*_0_ is computed using the next-generation matrix. The local and global stability of the disease-free equilibrium is analysed using the Jacobian matrix and Lyapunov function respectively. Furthermore, bifurcation analysis is performed using the Castillo-Chavez and Song theorem and sensitivity analysis is conducted on key parameters to identify their influence on disease dynamics. Numerical simulations of the model supports the analytical results. The findings of the study indicate that if the reproduction number is less than one, the solution converges to the disease-free state, signifying the asymptotic stability of the HPV-Cervical cancer free steady state. Crucially, the model demonstrates that increasing vaccination, screening and treatment rates significantly reduces HPV and cervical cancer incidence. This study underscores the value of mathematical modeling in informing the public health policy and provides a framework for optimizing control measures against HPV and Cervical cancer.

## Introduction

Cervical cancer is the fourth leading cause of cancer-related death in women after breast, lung, and colorectal cancers, with 341,834 deaths reported annually [1]. Almost 99% of cervical cancer cases are linked to infection with high risk human papillomavirus (HPV), an extremely common virus transmitted through sexual contact [1]. HPV was discovered to be the causative agent of cervical cancer in the 1970s by the Zur Hausen group [2]. It is estimated that the probability of a person being infected with human papillomavirus (HPV) in their lifetime reaches 70 to 80% [3]. HPV is a non-enveloped double-stranded DNA virus that can be classified into cutaneous or mucosal and can be either high-risk (hrHPV) or low-risk (lrHPV) [4]. Most of the HPV infections are asymptomatic and can disappear without treatment over the course of a few years. For instance, about 70% of HPV infections will regress spontaneously within a year and 90% within two years. However, in some people infection can persist for many years and can cause warts or low risk genotype of HPV, while other types lead to different kinds of cancers or high risk genotype of HPV including cervical cancer [5]. Persistent infection with high-risk HPV types can over time cause pre-cancerous cervical lesions that, if left untreated, progress to invasive cervical cancer [6]. Even though there are over 100 HPV genotypes, common low-risk HPV 6, 11, 42, 43, and 44 more often cause benign cutaneous lesions in the form of warts and only very rarely develop into tumors; by contrast, hrHPV 16, 18, 31, 33, 34, 35, 39, 45, 51, 52, 56, 58, 59, 66, 68, and 70 are associated with cervical neoplasia [7]. HPV-16 and HPV-18 are considered to contribute to about 70% of cervical cancer cases or precancerous lesions in women [8]. There is no specific treatment for HPV, but the visible symptoms or genital warts due to the disease can be treated, with the goal of reducing the spread of the infection [9]. Symptoms of early-stage cervical cancer may include irregular blood spotting or light bleeding between periods in women of reproductive age, postmenopausal spotting or bleeding, bleeding after sexual contact, and increased vaginal discharge, sometimes foul smelling. Vaccination is one of the most important primary intervention and equitable public health strategies in existence to combat infectious disease globally. HPV vaccination is given to stimulate the immune system to produce antibodies. Further, these antibodies will prevent HPV from infecting cervical cells. HPV vaccination include primary vaccination of pre pubertal boys and girls and secondary vaccination of women aged between 10 and 45 years. HPV vaccination can protect against the acquisition of new HPV infections; it does not reduce the progression of established cervical lesions [10]. Currently, three vaccines are administered worldwide for the prevention of HPV infection. Cervarix, Gardasil and Gardasil-9. Cervarix is a bivalent vaccine that prevents infection against high-risk types, HPV-16 and HPV-18, while Gardasil is a quadrivalent vaccine that prevents infection caused by both high-risk and low-risk types i.e HPV-16, HPV-18, HPV-6 and HPV-11 and Gardasil-9 is a nonavalent vaccine that protects against nine types of HPV which areHPV types, 6,11,16,18,31,33,45,52,58 [11].

Cervical cancer screening involves testing for HPV infection to detect pre-cancer and cancer among individuals who display no symptoms and may not even feel sick [12]. Cervical cancer screening remains an effective way for early detection of precancerous lesions and cancers. Screening can also detect cancer at an early stage where treatment has a high potential for cure [12]. World Health Organisation (WHO) has recommended three types of screening tests: HPV testing for high risk HPV types, visual inspection with Acetic Acid (VIA) as well as conventional (Pap) test and liquid-based cytology (LBC) [12]. Pap screening detects abnormal cervical cells, including pre-cancerous cervical lesionsand early cervical cancers [13], [14] and [15]. Pap cytology screening for the early detection of cervical neoplasia has been successful in reducing cervical cancer incidence and mortality [16]. Cervical cancer can be cured if diagnosed at an early stage and treated promptly. World Health Organisation (WHO) recommends the use of cryotherapy and Loop Electrosurgical Excision Procedure (LEEP) for treatment of pre-cancer lesions. Hysterectomy, Cone biopsy, Radical trachelectomy, Lymph node removal, chemoradiation,radiation therapy, surgery, clinical trials and radiation therapy are all used to treatment cervical cancer [17].

Mathematical modeling serves as a powerful and versatile tool across various disciplines, providing a structured approach to understanding, analyzing, and predicting complex phenomena. Several models have been proposed to study the effects of some factors on the transmission dynamics of infectious diseases and to provide guidelines as to how the spread can be controlled. In a study by [18], developed a mathematical modeling of cervical cancer with HPV transmission and vaccination. The study showed that vaccinating females can be one of the technique to reduce and control cervical cancer.

Research study by [19] have developed a mathematical model of cervical cancer with vaccination and treatment effectiveness. Authors concluded that vaccination in general, have an important role in reducing and controlling cervical cancer. In a study by [20], a mathematical model for HPV transmission with vaccination and screening was developed. They considered ten compartments; susceptible, vaccinated, infectious individuals without disease symptoms, infectious individuals with disease symptoms, individuals with persistent HPV infection, CIN1 symptomatic individuals, CIN2 symptomatic individuals, CIN3 symptomatic individuals, cancer-infected individuals and recovered individuals. The results showed that increasing the vaccination rate is the most effective way to reduce the basic reproduction number. In this study we propose a mathematical model to understand the transmission dynamics of HPV with Cervical cancer infection in the population. Our study extends and improve the work [20] which takes into account vaccination and screening. In addition, we will introduce treatment compartment as a core intervention alongside vaccination and screening. Our model also accounts for individuals who are infected with HPV but remain unaware of their infection and subsequently develop cervical cancer.

## Model Formulation

A model for HPV and cervical cancer with screening, vaccination and treatment is formulated by subdividing the total population at time t, denoted by *N* (*t*), into seven compartmental states. Susceptible individuals at time t denoted by *S*(*t*), Individuals vaccinated against HPV at time t denoted by *V* (*t*), Individuals infected with HPV but are unaware denoted by *I*_*u*_(*t*), Individuals screened for precancerous lesions and cervical cancer at time t denoted by *IS*_*c*_(*t*), Individuals infected with cervical cancer denoted by *C*(*t*), treated individuals at time t denoted by *T* (*t*) and recovered individuals at time t denoted by *R*(*T* ). With this subdivision the total population *N* (*t*) is governed by,

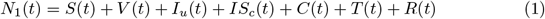

### Model Assumptions

1. Homogeneous mixing within each compartment, meaning that all individuals have an equal probability of interacting with each other.
2. Parameters vary from compartment to compartment but are identical for all individuals in a given compartment.
3. The standard incidence principle is applied to model the transmission dynamics of cervical cancer. This principle assumes that the transmission rate of infection depends on proportion of infected individuals rather than absolute numbers.
4. In all compartments, the natural death rate is *µ*.
5. The HPV force of infection *λ*_**1**_ is given as

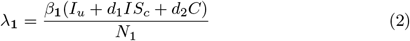

Where *β*_**1**_ is the effectual contact rate of the HPV.

The rate of vaccination against HPV is *θ*. Vaccinated individuals can also become infected with HPV when vaccine wanes. When the vaccine efficacy is 100%, the vaccinated individuals cannot become infected.

The HPV unaware infected individuals are screened and join the screened infected class at a rate *ρ*. Some of the unaware infected individuals progress to cervical cancer at rate *τ* and others recover naturally through body immune system at rate *ϵ*. The screened infected individuals found with precancerous lesions are treated at rate *γ* and move to recovery at rate *ϕ*. Infected individuals that are screened and found with cervical cancer, move to cervical cancer class at rate *ξ* and seeks treatment at rate *α*. The recovered individuals lose immunity at the rate of *ω* joins the susceptible class.

Individuals infected with cervical cancer suffer disease-induced death at the rates *δ*_**1**_. In this section using parameters described in Table 1, and the given assumptions the Flow chart of the HPV-Cervical cancer infection is given by Fig 1.

**Table 1.** Description of model parameters.

| Parameter | Description |
| --- | --- |
| $\Lambda$ | Recruitment rate to the population |
| $\theta$ | HPV vaccination rate |
| $\theta_1$ | Vaccine waning |
| $\beta_1$ | HPV transmission rate |
| $\epsilon$ | Immune clearance rate of HPV infection |
| $\tau$ | Progression rate from HPV to cervical cancer |
| $\gamma$ | Treatment rate for screened HPV-positive individuals |
| $\alpha$ | Treatment rate for cervical cancer patients |
| $\phi$ | Recovery rate for treated individuals |
| $\omega$ | Rate of return from recovered to susceptible |
| $\rho$ | Screening rate for HPV-infected individuals |
| $\xi$ | Detection rate of cervical cancer in screened individuals |
| $d_1$ | Transmission reduction factor for screened individuals |
| $d_2$ | Transmission reduction factor for cancer patients |
| $\mu$ | Natural death rate |
| $\delta_1$ | Cervical cancer-induced mortality rate |

**Fig 1.**
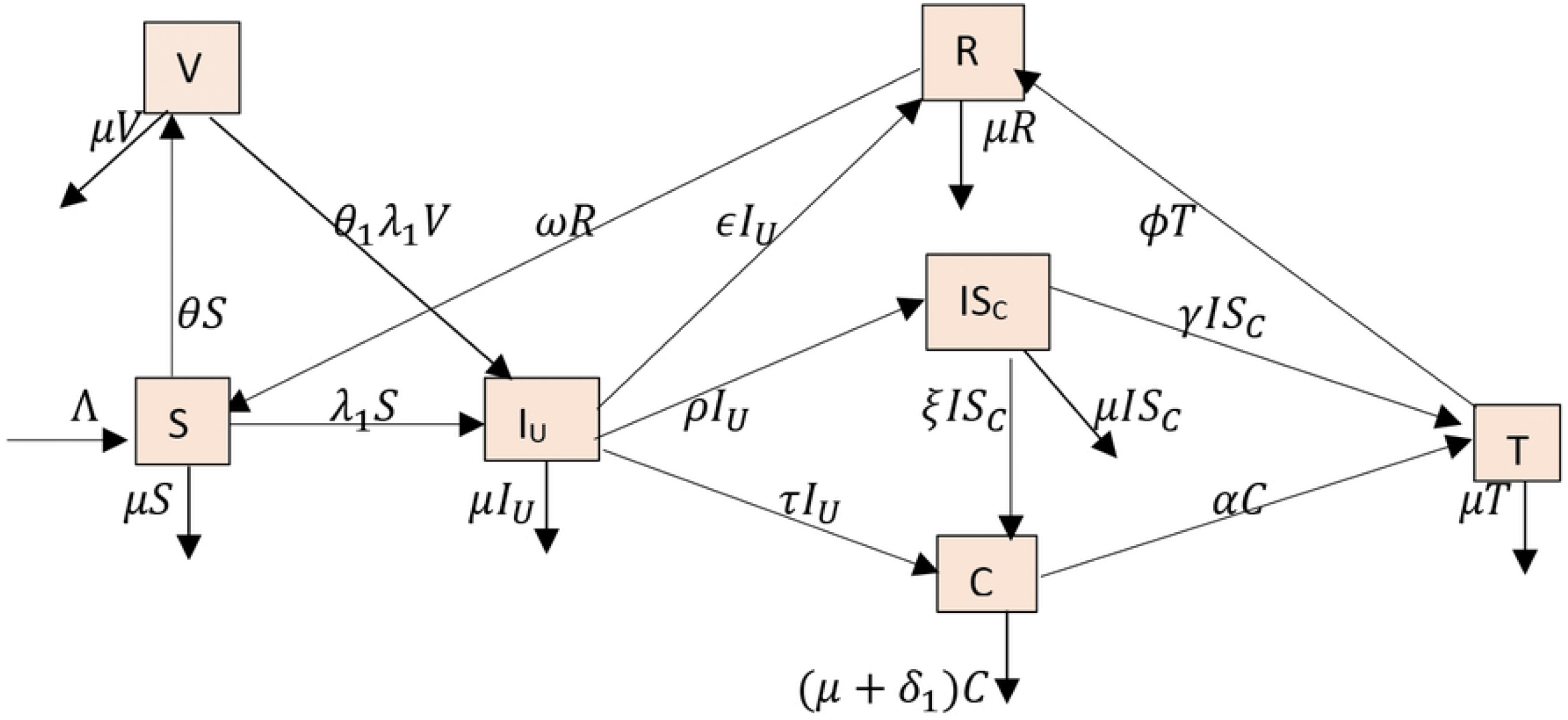
Flow chart of HPV-Cervical cancer infection.

**Fig 2.**
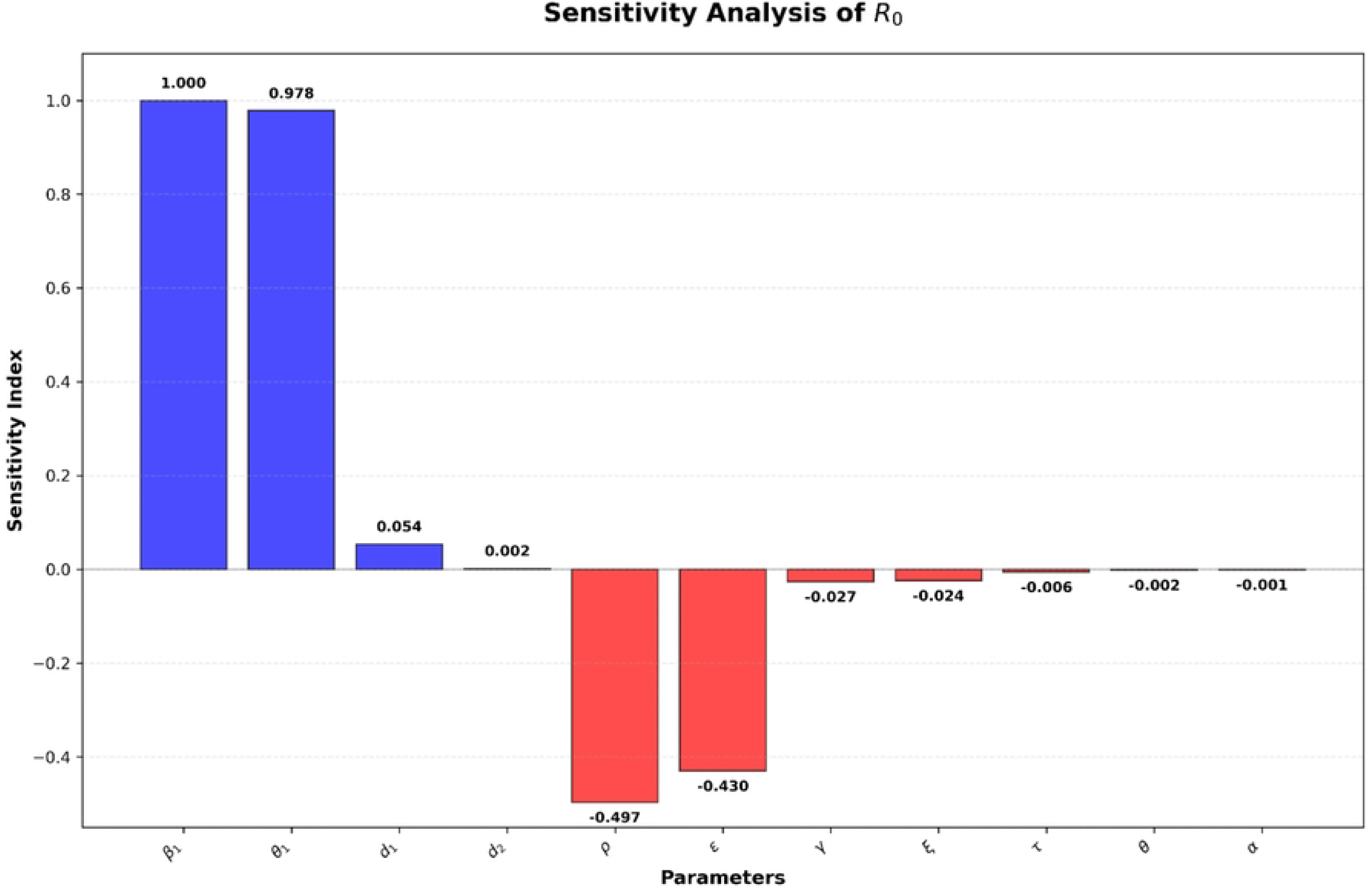
Sensitivity Indices diagram for *R*_*oc*_.

### Model Equations

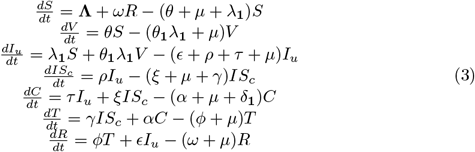

with initial conditions

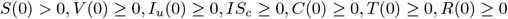

## Model Analysis

The model (3) is both biologically and mathematically meaningful if and only if all the model solutions are non-negative and bounded in the invariant region, t*>*0.

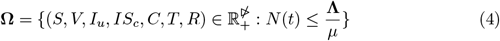

### Theorem 0.1.

(***Positivity of Solutions:***)

*Given that the initial conditions of system (3) are S*(0) *>* 0, *V* (0) ≥ 0, *I*_*u*_ ≥ 0, *IS*_*c*_ ≥ 0, *C*(0) ≥ 0, *T* (0) ≥ 0, *and R*(0) ≥ 0, *the solutions S*(*t*), *V* (*t*), *I*_*u*_(*t*), *IS*_*c*_(*t*), *C*(*t*), *T* (*t*), *R*(*t*) *are non negative for all t>0*.

*Proof*.

Assume that

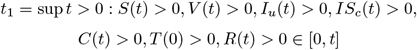

Thus *t*_1_ *>* 0

Considering the first equation of the model (3), we obtain

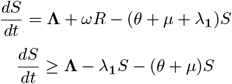

We use the integrating factor method to solve the equation

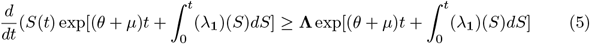

Integrating both sides yields

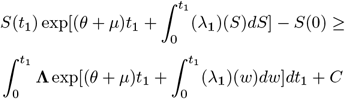

Where C is the constant of integration. Hence

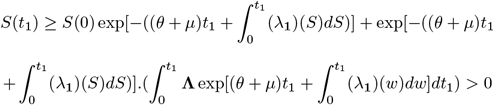

It is clear from the inequality above that

*S*(*t*_1_) ≥ 0 is non-negative

From the second equation in system (3), we obtain

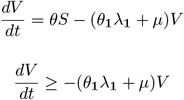

Using the integrating factor method to solve inequality, we have

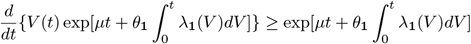

Integrating both sides yields

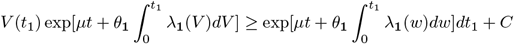

Hence,

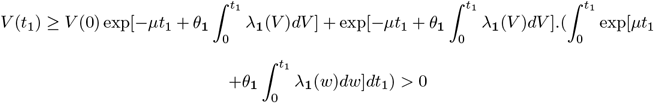

Hence, *V* (*t*_1_) *>* 0∀*t*_1_ *>* 0

Similarly, it can be shown that

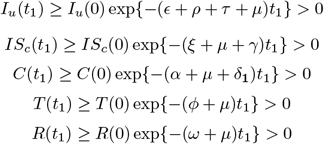

### Theorem 0.2.

*(**The invariant region:**)*

*All the feasible non-negative solutions of the HPV-Cervical cancer model (3) with initial conditions S(0) > 0, V (0)*≥*0, Iu*≥ *0, ISc*≥ *0,C(0)*≥ *0, T(0)*≥ *0, and R(0)*≥ *0, are bounded in the region (4)*.

*Proof*.

By adding up the right-hand sides of the HPV-Cervical cancer model (3), the total population is given by

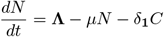

In absence of mortality due to cervical cancer, the above becomes

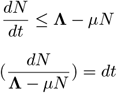

Taking integrals of both sides

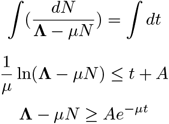

Where A is constant. Applying the initial conditions, we obtain the relation,

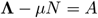

Therefore, **Λ** − *µN* ≥ (**Λ** − *µN* (0))*e*^−*µt*^

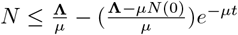 As t → ∞, the population size 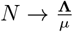.

This implies that 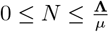 and 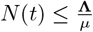. Also if 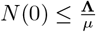, then 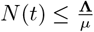.

Therefore, N(t) is bounded above. Subsequently, *S*(*t*), *V* (*t*), *I*_*u*_(*t*), *IS*_*c*_(*t*), *C*(*t*), *T* (*t*)*andR*(*t*)are bounded above. Thus, in **Ω**, system (3) is well posed. Hence, it is sufficient to study the dynamics of the system in **Ω**.

### Local stability of the model disease-free equilibrium point (DFE)

In this section, we obtain the equilibrium point at which HPV-cervical cancer is eradicated from the population. Letting the right hand side of equation(3) to zero and letting *I*_*u*_ = *IS*_*c*_ = *C* = 0 leads to,

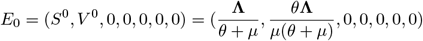

The basic reproduction number of cervical cancer sub-model is defined as the average number of secondary infections caused by a single infected individual in a susceptible population. It can be obtained by using the approach of the next-generation matrix as given in [21]. The next-generation matrix is defined as: *K* = *FV* ^−1^ and *R*_0_ = *ρFV* ^−1^ where *ρFV* ^−1^ denotes the spectral radius of *FV* ^−1^, F is the matrix of new infection terms and V is the matrix of transition terms. Using the Next Generation Matrix, we consider only the infectious classes in the system of differential equations in (3) :

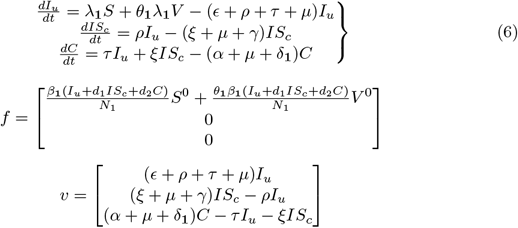

Then Jacobian matrix of *f* and *v* are obtained by F and V at disease-free equilibrium:

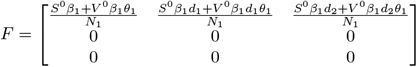

Where 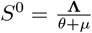 and 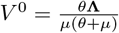

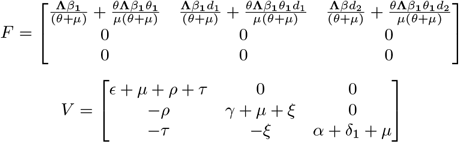

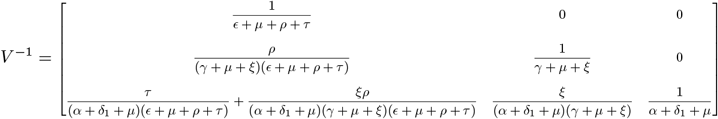

By computing the eigenvalues of *FV* ^−1^ and selecting the dominant eigenvalues,

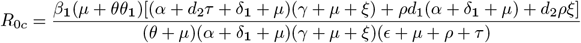

#### Theorem 0.3.

*The disease-free equilibrium point of the HPV-Cervical cancer model given in Eq (3) is locally asymptotically stable whenever R*_0*c*_ *<* 1 *is and unstable otherwise*.

*Proof*.

The local stability of the disease-free equilibrium point of HPV-Cervical cancer infection model (3) is studied by applying the Routh-Hurwitz stability criteria. The Jacobian matrix evaluated at *E*_0_ is obtained as

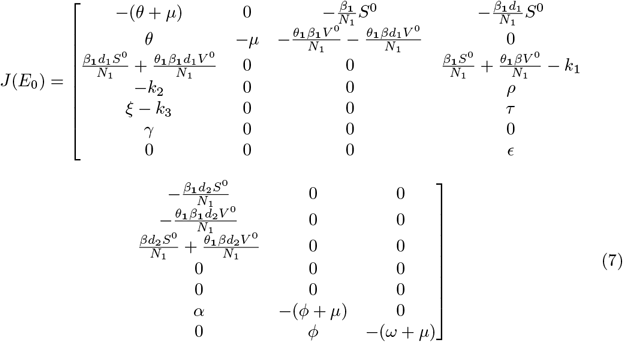

where

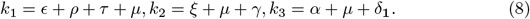

To obtain the eigenvalue of (7),

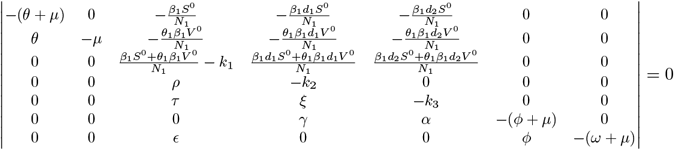

The characteristic polynomial of 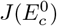 is given by

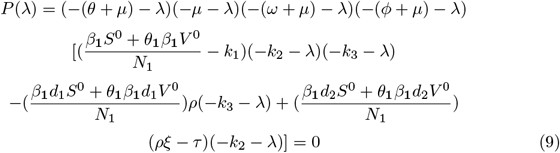

Simplifying the equation(9), we have

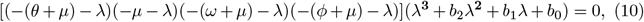

where

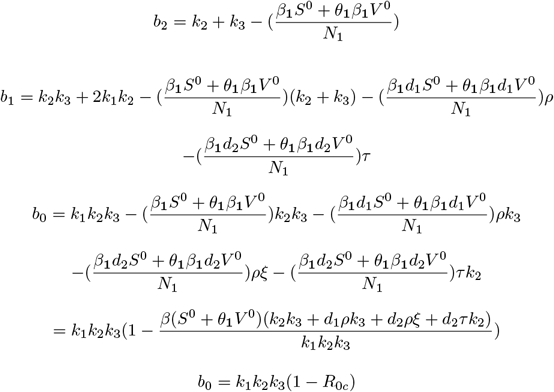

The following eigenvalues with negative real parts can be obtained from the polynomial *λ*_**1**_ = −(*θ* + *µ*), *λ*_**2**_ = −*µ, λ*_**3**_ = −(*ω* + *µ*), *λ*_**4**_ = −(*ϕ* + *µ*)

Because all model parameters are positive, *a >* 0 and *b >* 0 if and only if *R*_0*c*_ *<* 1. Therefore, by Routh - Hurwitz criterion, all roots of the characteristic polynomial Eq (9) have negative real part, if *R*_0*c*_ *<* 1. We therefore conclude that the disease - free equilibrium 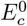 is locally asymptotically stable whenever *R*_0*c*_ *<* 1. The biological implication of proposition (0.3) is that if *R*_0*c*_ *<* 1, HPV-Cervical cancer will be eliminated from the model population.

#### Theorem 0.4.

*The disease-free equilibrium of the system (3) is globally asymptotically stable if R*_0*c*_ *<* 1 *and unstable if R*_0*c*_ *>* 1.

*Proof*.

Define the Lyapunov function

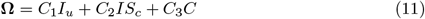

On differentiating **Ω** with respect to t in (11), we have

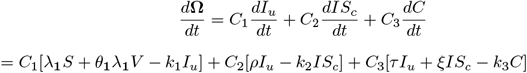

where 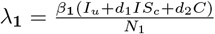, *k*_1_, *k*_2_ and *k*_3_ are given in (8)

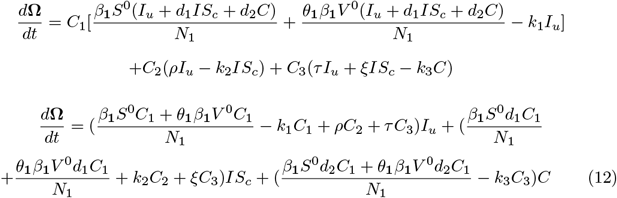

Given that

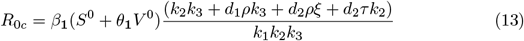

From equation (12) and using (13)

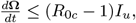

where 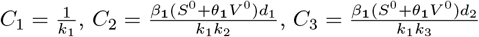

Because all model parameters are non-negative, it follows that 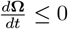 for *R*_0*c*_ ≤ 0, with 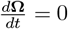 if and only if *I*_*u*_ = *IS*_*c*_ = *C* = 0. Substituting *I*_*u*_ = 0, *IS*_*c*_ = 0 and *C* = 0 in (3) shows that 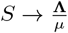 as *t* → ∞. Hence **Ω** is a Lyapunov function on **Ω**_**c**_ and the largest compact invariant set in 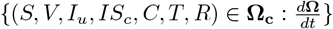 is 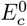. Thus by LaSalle’s Invariance Principle [21], every solution of (3) with initial condition in **Ω**_**c**_ approaches 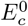, as *t* → ∞ whenever *R*_0*c*_ ≤ 1.

The endemic equilibrium points components are obtained from solving for 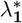 in the polynomial (20) and substituting the positive values of 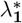 into the expression in Eq (14). Furthermore, it follows from Eq (21) that the coefficient A is always positive, and E is positive(negative) if *R*_0*c*_ is less(greater) than one. From Theorem 2 of Van den and Watmough [21], the following results follows.

#### Theorem 0.5.

*The unique endemic equilibrium* 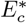 *of the HPV-Cervical cancer model (3) is locally asymptotically stable if R*_0*c*_ *>* 1 *and unstable otherwise*.

### Existence of the endemic equilibrium point

The HPV-Cervical cancer infection endemic equilibrium is given by 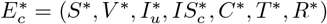: which was obtained by setting the right hand side of equation (3) to zero.

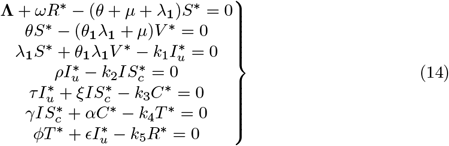

Where 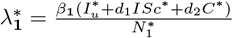, *k*_1_, *k*_2_ and *k*_3_ are given in (8), *k*_4_ = (*ϕ* + *µ*), *k*_5_ = (*ω* + *µ*) and *k*_6_ = (*θ* + *µ*)

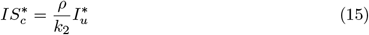

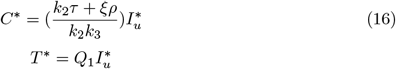

where

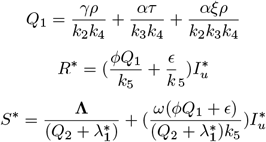

Where

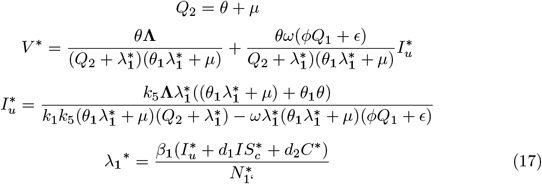

We get,

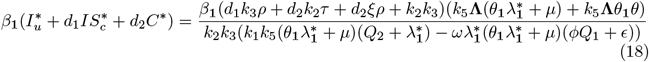

Where,

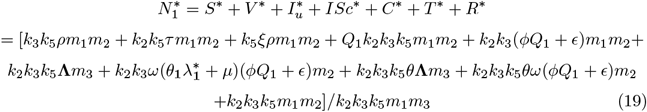

Where,

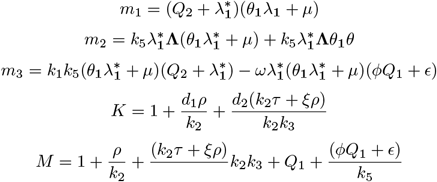

Putting (18) and (19) in (17) at 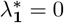, we get the following polynomial.

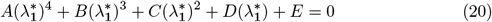

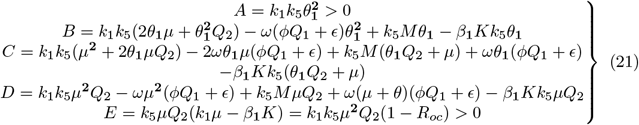

The endemic equilibrium points components are obtained from solving for 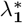 in the polynomial (20) and substituting the positive values of 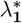 into the expression in (14). Furthermore, it follows from (21) that the coefficient A is always positive, and E is positive(negative) if *R*_0*c*_ is less(greater) than one. From Theorem 2 of Van den and Watmough [21], the following results follows.

#### Theorem 0.6.

*The sub-model (3) has*

i. *four or two endemic equilibria if B >* 0, *C >* 0, *D >* 0 *and* ℛ_0*c*_ *<* 1;
ii. *two endemic equilibria if B >* 0, *C >* 0, *D <* 0 *and* ℛ_0*c*_ *<* 1;
iii. *no endemic equilibrium otherwise, if* ℛ_0*c*_ *<* 1.

Item *(i)-(ii)* of Theorem 6 illustrates the condition for the phenomenon of backward bifurcation in the cervical cancer model (3) i.e., a local asymptotic stable disease-free equilibrium point co-exists with a local asymptotic stable endemic equilibrium point whenever *R*_0*c*_ *<* 1. The epidemiological consequence is that the classical epidemiological requirement of having the reproduction number *R*_0*c*_ to be less than one, even though necessary is not sufficient for the effective control of the HPV and cervical cancer disease transmission. The existence of the backward bifurcation phenomenon in the sub model is now explored.

#### Bifurcation analysis

It is important to note that the disease-free equilibrium interchanges its stability with the endemic equilibrium as the threshold parameter *R*_0*c*_ is varied, we wish to investigate the direction of the bifurcation (forward/transcritical bifurcation or backward/subcritical bifurcation). Global stability of the model equilibria implies the model does not exhibit the phenomena of backward bifurcation where co-existence of both a stable DFE with a stable endemic equilibrium cannot hold [22]. We use the centre manifold theory as described in Theorem 4.1 from [23]. To apply this theory, the following simplification and change of variables are made first of all.

Let *x* = (*x*_1_, *x*_2_, *x*_3_, *x*_4_, *x*_5_, *x*_6_, *x*_7_)^*T*^ = (S, V, *I*_*u*_, *IS*_*c*_, *C*, T, *R*)^*T*^, the cervical cancer sub-model can be rewritten in the form 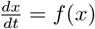, where *f* (*x*), = (*f*_1_(*x*), *f*_2_(*x*), *f*_3_(*x*), *f*_4_(*x*), *f*_5_(*x*), *f*_6_(*x*), *f*_7_(*x*)) as follows

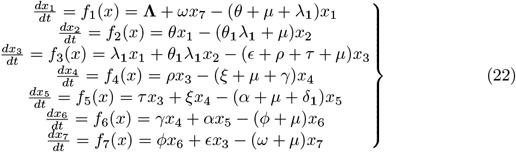

Where

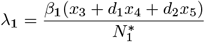

and 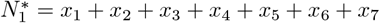

Choosing 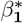 as the bifurcation parameter and setting *R*_0*c*_ = 1, we obtain

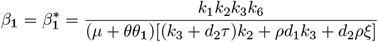

The Jacobian matrix of the system (22) evaluated at 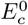 and 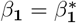 is given by

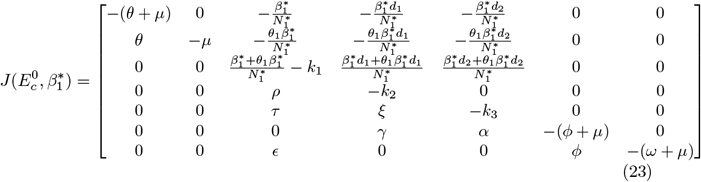

The eigenvalues of matrix 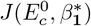 are *λ*_**1**_ = − (*θ* + *µ*), *λ*_**2**_ = − *µ, λ*_**3**_ = − (*ω* + *µ*), *λ*_**4**_ = − (*ϕ* + *µ*), *λ*_**5**_ = − *k*_3_. The remaining eigenvalues are the roots of the characteristic equation

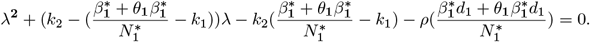

which are *λ*_6_ = 0 and 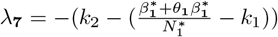. Thus, 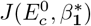 has simple zero eigenvalue and other eigenvalues have negative sign. Hence 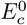 is a non-hyperbolic equilibrium, when 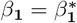

Next, we calculate the right and left eigenvectors associated to the zero eigenvalue. let *w* = (*w*_1_, *w*_2_, *w*_3_, *w*_4_, *w*_5_, *w*_6_, *w*_7_)^*T*^ be the right eigen vector and can be associated with simple zero eigenvalue and can be obtained by solving equation 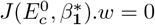.

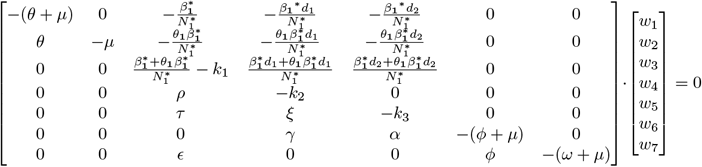

Thus we have:

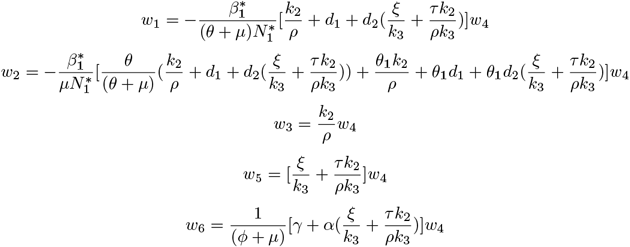

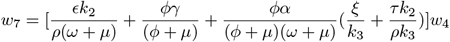

Also, let *v* = (*v*_1_, *v*_2_, *v*_3_, *v*_4_, *v*_5_, *v*_6_, *v*_7_) be the left eigenvector corresponding to simple zero eigenvalues, obtained by setting and solving 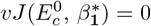. This gives, *v*_1_ = 0, *v*_2_ = 0, *v*_6_ = 0, *v*_7_ = 0, 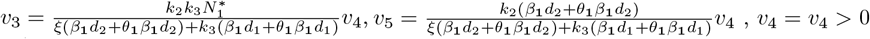

In order that the required conditions *v*.*w* = 1, the product gives

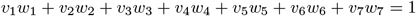

Again substituting the corresponding components in the preceding equation, we have

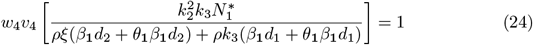

The Eq(24) is satisfied if we choose *w*_4_ = *ρξ*(*β*_**1**_*d*_2_ + *θ*_**1**_*β*_**1**_*d*_2_) + *ρk*_3_(*β*_**1**_*d*_1_ + *θ*_**1**_*β*_**1**_*d*_1_) and 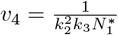

*Bifurcation constants a and b:* The bifurcation coefficients a and b at the DFE *E*^0^ is given by

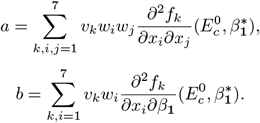

Since *v*_1_ = *v*_2_ = *v*_5_ = *v*_6_ = *v*_7_ = 0, we do not need the derivatives of *f*_1_, *f*_2_, *f*_5_, *f*_6_ and *f*_7_. Also, the second order partial derivatives of *f*_4_ are all zero. Thus we need only the second order partial derivatives of *f*_3_ where

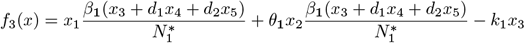

Therefore, the nonzero second order partial derivatives of *f*_3_ at 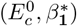 are

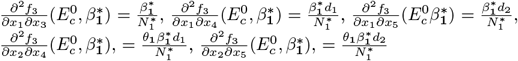

The bifurcation coefficient *a* at the DFE 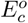 is given by

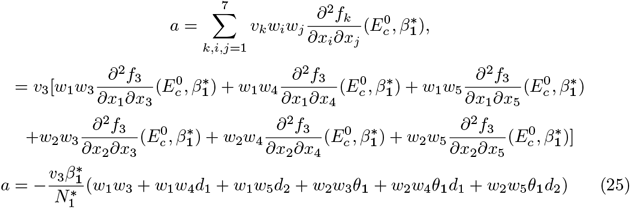

The second bifurcation coefficient *b* is given by

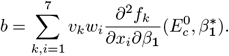

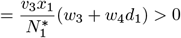

From the computations, we have *a <* 0 and *b >* 0 at 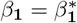. Since the bifurcation coefficient b is positive, it follows that, Theorem 4.1 in [23] that the model (3) will undergo backward bifurcation if the coefficient, a, given by Eq (25) is positive. Setting the HPV re-infection term *ω* and vaccine waning *θ*_**1**_ to zero, it is observed that the bifurcation coefficient, *a <* 0. Hence, backward bifurcation does not occur in the HPV-Cervical cancer model, in the absence of HPV re-infection and vaccine waning. The epidemiological interpretation is that if recovery from HPV does not confer lifelong immunity and there is no 100% vaccine efficacy, then the control of HPV-Cervical cancer becomes difficult even when the associated reproduction number *R*_0*c*_ *<* 1.

### Sensitivity Analysis

Sensitivity analysis is carried out to determine the parameters that enhances the spread as well as control of an infection in a population. Sensitivity analysis in mathematical modeling involves studying how changes in the input parameters of a model affect the output or the solutions. It helps to understand the robustness and reliability of a model by assessing the impact of variations in its input values. In this section, we followed the approach of [24] and [25]. The most sensitive parameter is the one with the highest magnitude as compared to others.

The sensitivity index of *R*_0*c*_ with respect to p; where *p* ∈ (*β*_**1**_, *θ*_**1**_, *τ, ξ, ρ, θ, α, γ, ϵ, d*_1_, *d*_2_) are computed at Table 2.

**Table 2.** Sensitivity indices of *R*_0_.

| Parameter | Sensitivity index |
| --- | --- |
| $\beta_1$ | +1.0 |
| $\theta_1$ | +0.978 |
| $\tau$ | -0.00609 |
| $\xi$ | -0.0241 |
| $\rho$ | -0.497 |
| $\theta$ | -0.00216 |
| $\alpha$ | -0.00116 |
| $\gamma$ | -0.0266 |
| $\epsilon$ | -0.430 |
| $d_1$ | +0.0537 |
| $d_2$ | +0.00167 |

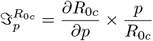

From Table 2 we observe that the parameters *τ, ξ, ρ, θ, α, γ, ϵ* have negative signs. This means an increase (decrease) of these parameters, then the value *R*_0*c*_ will decrease (increase). The remaining parameter indices *β*_**1**_, *θ*_**1**_, *d*_1_, *d*_2_ a have a positive sign and this indicates a decrease (increase) in these parameters and decrease (increase) in the reproduction number. From Table 2, it shows that the most sensitive parameters is transmission rate *β*_**1**_. From Table 3, If the screening rate *ρ* varies, the sensitivity index of the parameter *ρ* varies which shows that the screening rate is the most sensitive parameter. If screening rate is increased by 30%, the *R*_0*c*_ reduces by 27%. If the vaccination rate *θ* varies, the sensitivity index of the parameter *θ* varies which shows that vaccination rate *θ* is the most sensitive parameter. If vaccination rate is increased by 35% the reproduction number is reduced by 15%. If treatment effectiveness is increased by 30%, the reproduction number is reduced by 9%

**Table 3.** Sensitivity indices of *R*_0_ for varying screening, vaccination and treatment.

| Parameter | Varying Parameter value | Sensitivity Index value |
| --- | --- | --- |
| $\rho$ | for $\rho=0.3$ | -0.328 |
| $\rho$ | for $\rho=0.6$ | -0.428 |
| $\rho$ | for $\rho=0.9$ | -0.497 |
| $\rho$ | for $\rho=1.2$ | -0.534 |
| $\theta$ | for $\theta=0.2$ | -0.034 |
| $\theta$ | for $\theta=0.5$ | -0.128 |
| $\theta$ | for $\theta=0.8$ | -0.216 |
| $\theta$ | for $\theta=0.9$ | -0.257 |
| $\tau$ | for $\tau=0.05$ | -0.029 |
| $\tau$ | for $\tau=0.1$ | -0.057 |
| $\tau$ | for $\tau=0.2$ | -0.109 |

### Numerical simulations

Numerical simulation in mathematical modeling involves using computational techniques to approximate solutions to mathematical models. This approach is particularly useful when analytical solutions are difficult or impossible to obtain. From the model parameters we have obtained the values used to draw the following graphs. The initial conditions used for the graphs were as follows; S(0)=50000, V(0)=1000, *I*_*u*_(0)=10000, *IS*_*c*_=1000, C(0)=5, T(0)=500, R(0)=0. The graphs were drawn using python software which utilized Rungekutta order four method. The parameter values used and their corresponding references sources are summarized in Table 4.

**Table 4.** The parameter values of the model.

| Parameter | Value(per year) | Reference |
| --- | --- | --- |
| $\Lambda_1$ | 288802 | [19] |
| $\theta$ | 0.8 | [19] |
| $\theta_1$ | 0.9 | [18], [26] |
| $\beta_1$ | 1.5 | Assumed |
| $\epsilon$ | 0.7 | [27] |
| $\rho$ | 0.9 | [22] |
| $\tau$ | 0.01 | Assumed |
| $\xi$ | 0.15 | [22] |
| $\gamma$ | 0.1 | [27] |
| $\alpha$ | 0.85 | [28] |
| $d_1$ | 0.2 | Assumed |
| $d_2$ | 0.01 | Assumed |
| $\delta_1$ | 0.8 | [27] |
| $\mu$ | 0.0162 | [18] |
| $\phi$ | 0.0576 | [19] |
| $\omega$ | 0.3 | [20] |

From Fig 3 it shows evolution of all population compartment (susceptible, vaccinated, HPV infected, screened, Cervical cancer, treated, recovered) over time without coordinated interventions. The susceptible population decreases,steadily as individuals move into the infected or vaccinated compartment. The HPV-infected compartment peaks at approximately 15% of the population before declining primarily due to natural recovery and the progression of a fraction of individuals to cervical cancer. This demonstrates the natural history of the disease, where significant cancer incidences emerges from the population of untreated HPV infections. In contrast, Fig 4 demonstrates the shift of the disease when both vaccination and screening are implemented and without treatment, *τ* = 0. The vaccinated compartment rapidly expands and stabilizes at high level. Cervical cancer cases accumulate over time which represent a failure of secondary prevention.

**Fig 3.**
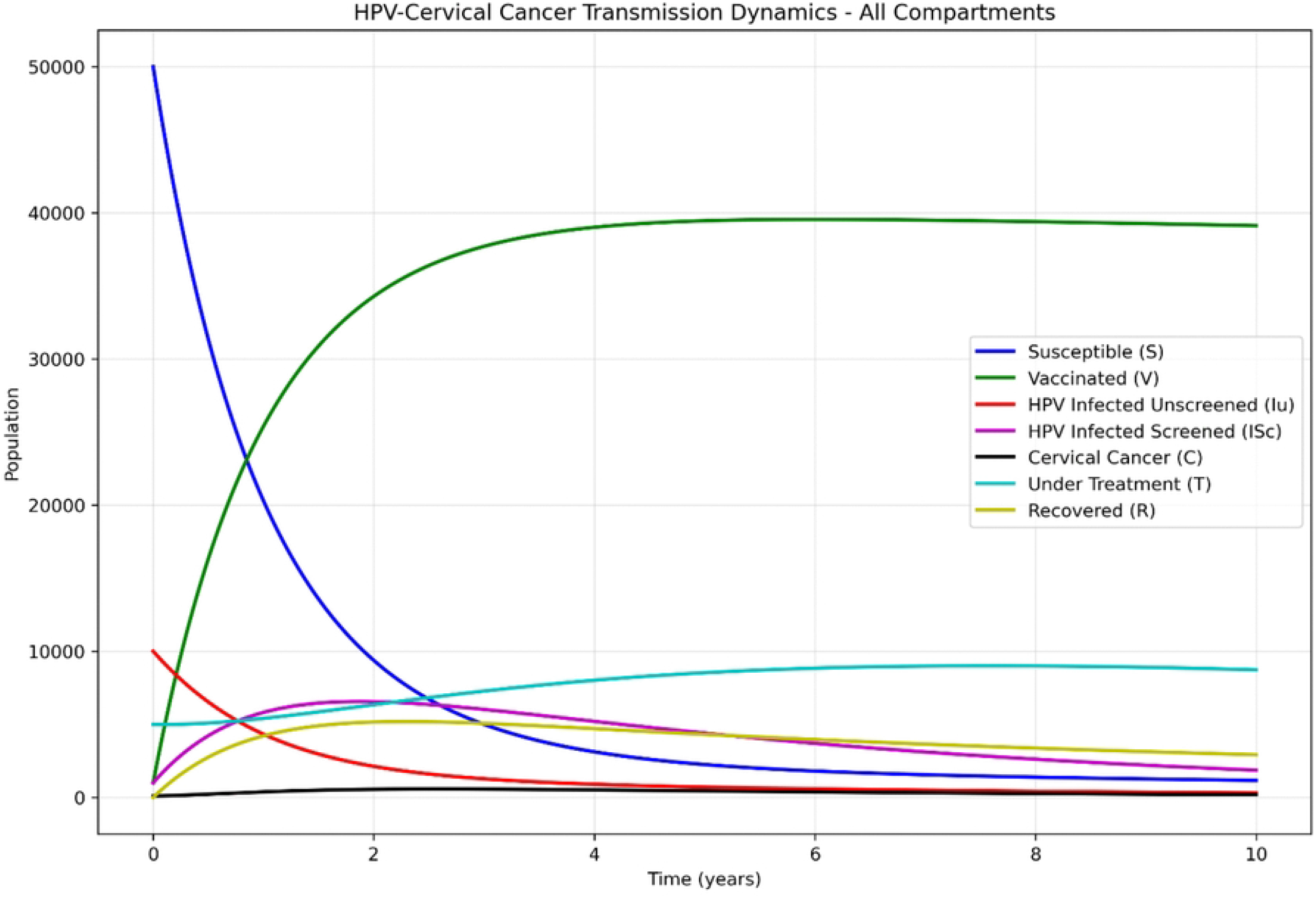
HPV-Cervical cancer model solution dynamics.

**Fig 4.**
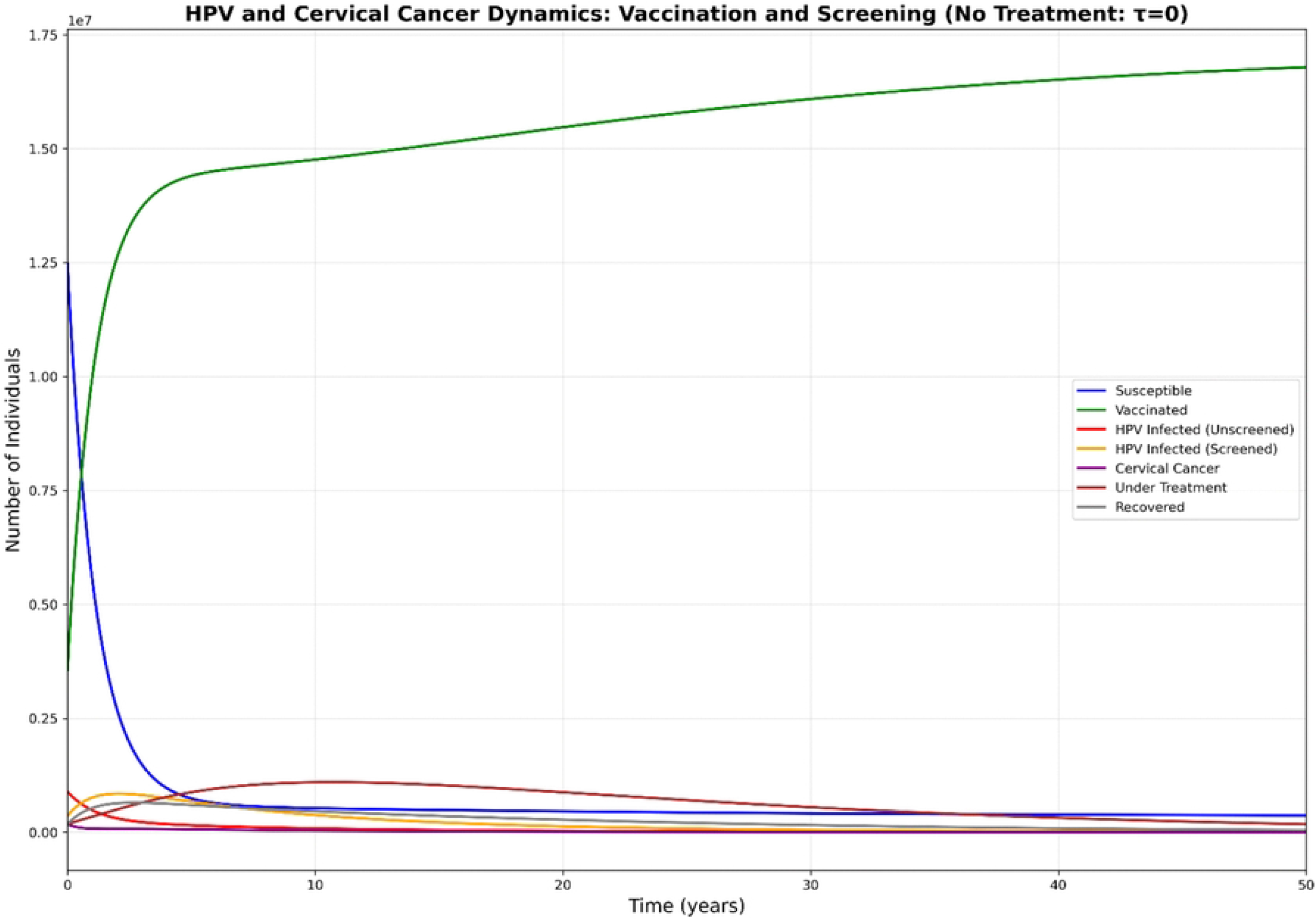
HPV and Cervical cancer dynamics without treatment.

Fig 5 provide a direct comparison of interventions on HPV prevalence and cervical cancer incidence. From Fig 6 and Fig 7, it shows vaccination has a stronger impact on reducing HPV prevalence because it acts primarily on the force of infection preventing new cases. Also screening alone shows modest reduction in total HPV incidence because screening detects existing infections but does not prevent new ones. Combined intervention give the best outcome for HPV and Cervical cancer control because vaccination reduces new infections while screening manages the existing ones.

**Fig 5.**
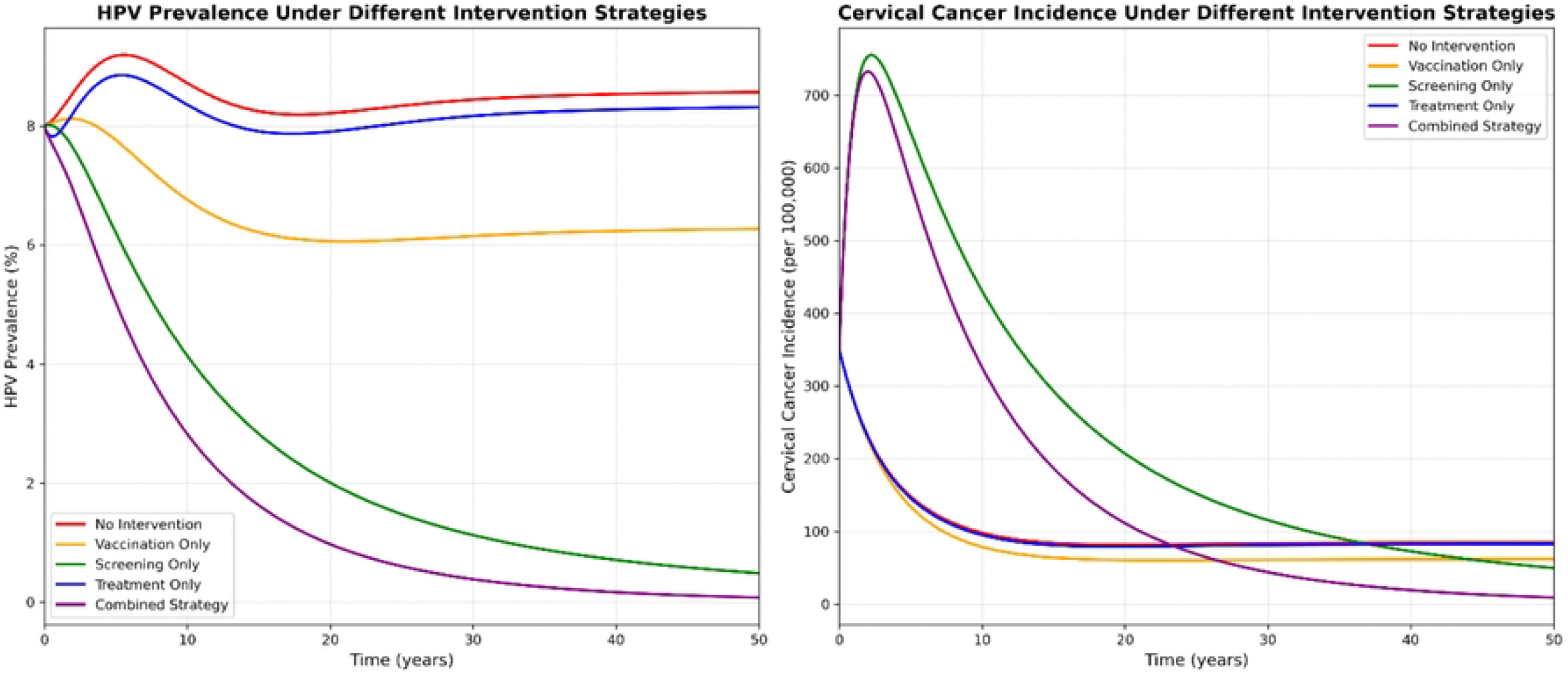
Comparative Impact of all interventions.

**Fig 6.**
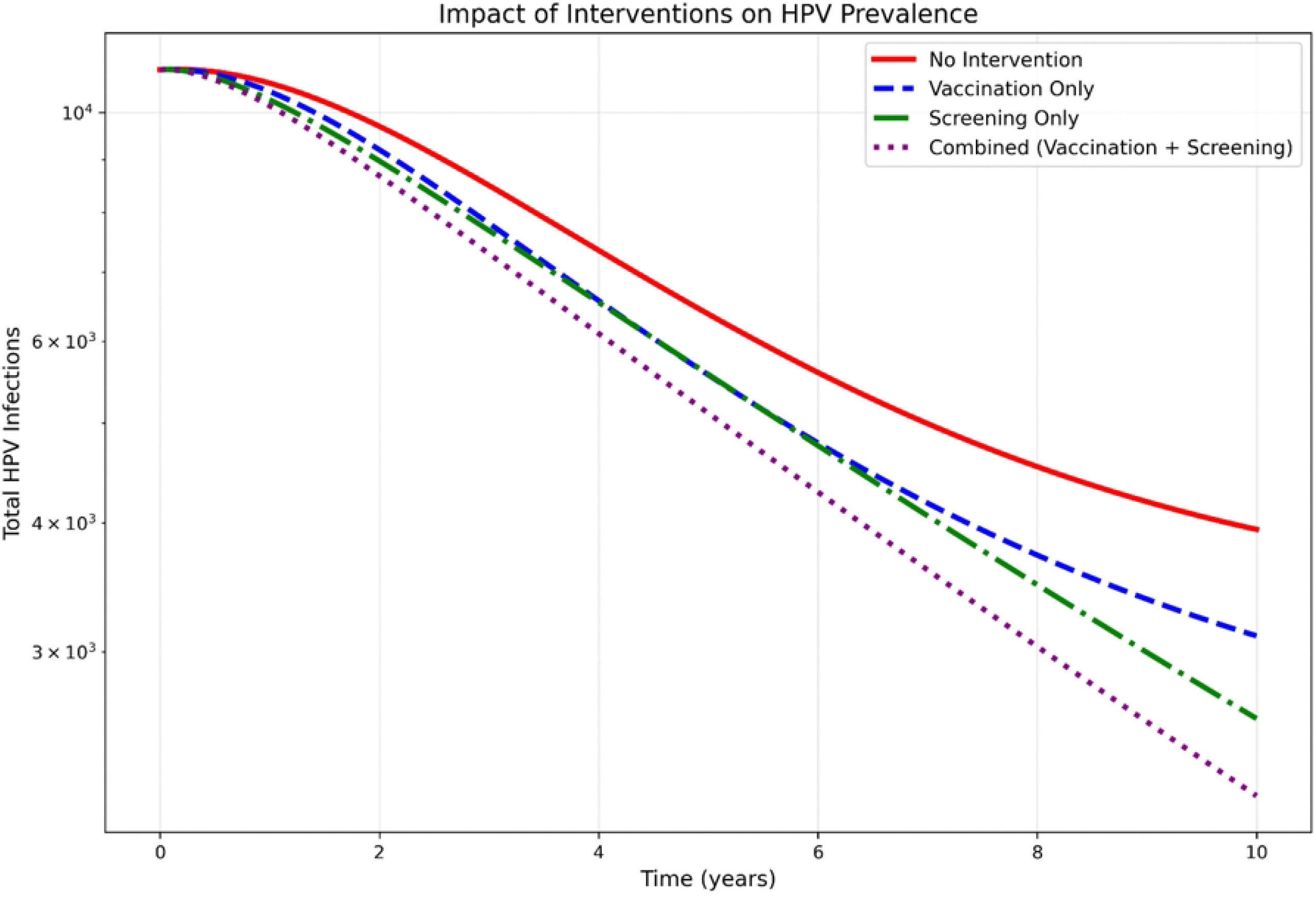
Impact of screening and vaccination on HPV prevalence.

**Fig 7.**
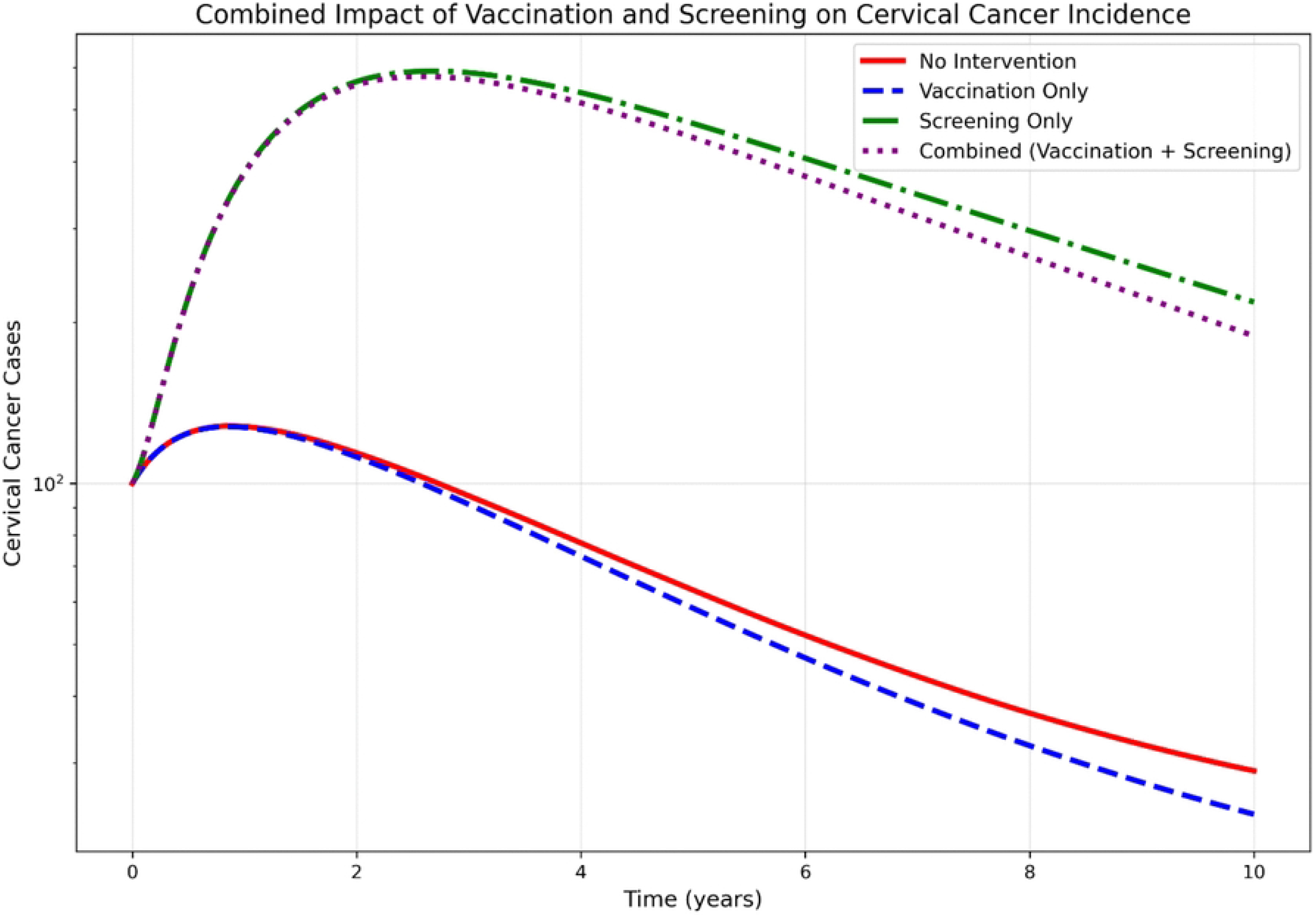
Impact of screening and vaccination on cervical cancer.

Fig 8 and Fig 9 shows effects of varying screening rates on HPV and Cervical cancer incidences. Screening moves people from *I*_*u*_(unscreened) to *IS*_*c*_(screened) but they are still infected. Cancer prevention comes from early treatment of pre-cancerous lesions . HPV transmission continues because screened individuals can still transmit though at reduced rate *d*_1_. This shows that screening is a powerful tool for cervical cancer control but is insufficient alone for controlling overall HPV transmission.

**Fig 8.**
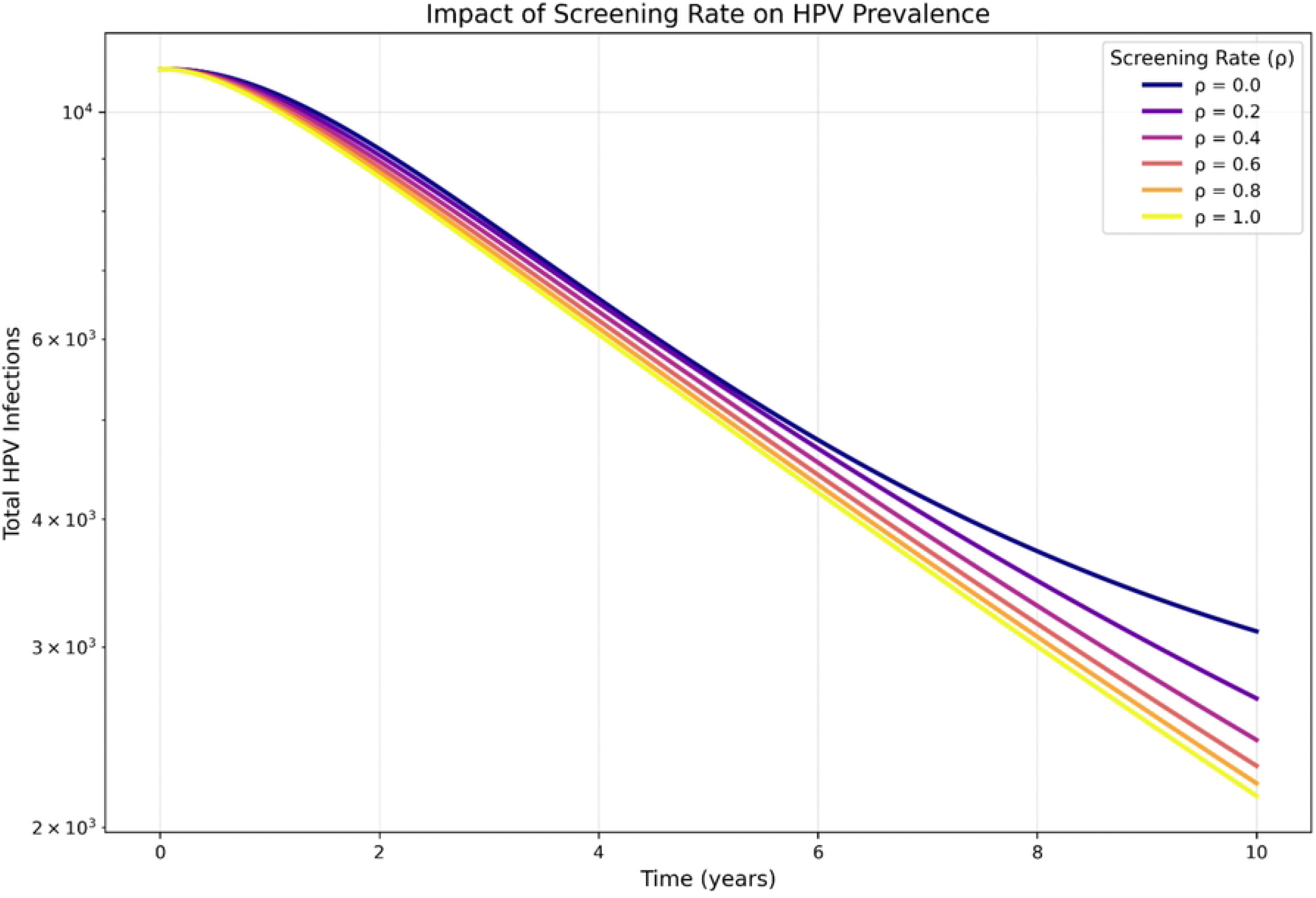
Varying screening rates on HPV prevalence.

**Fig 9.**
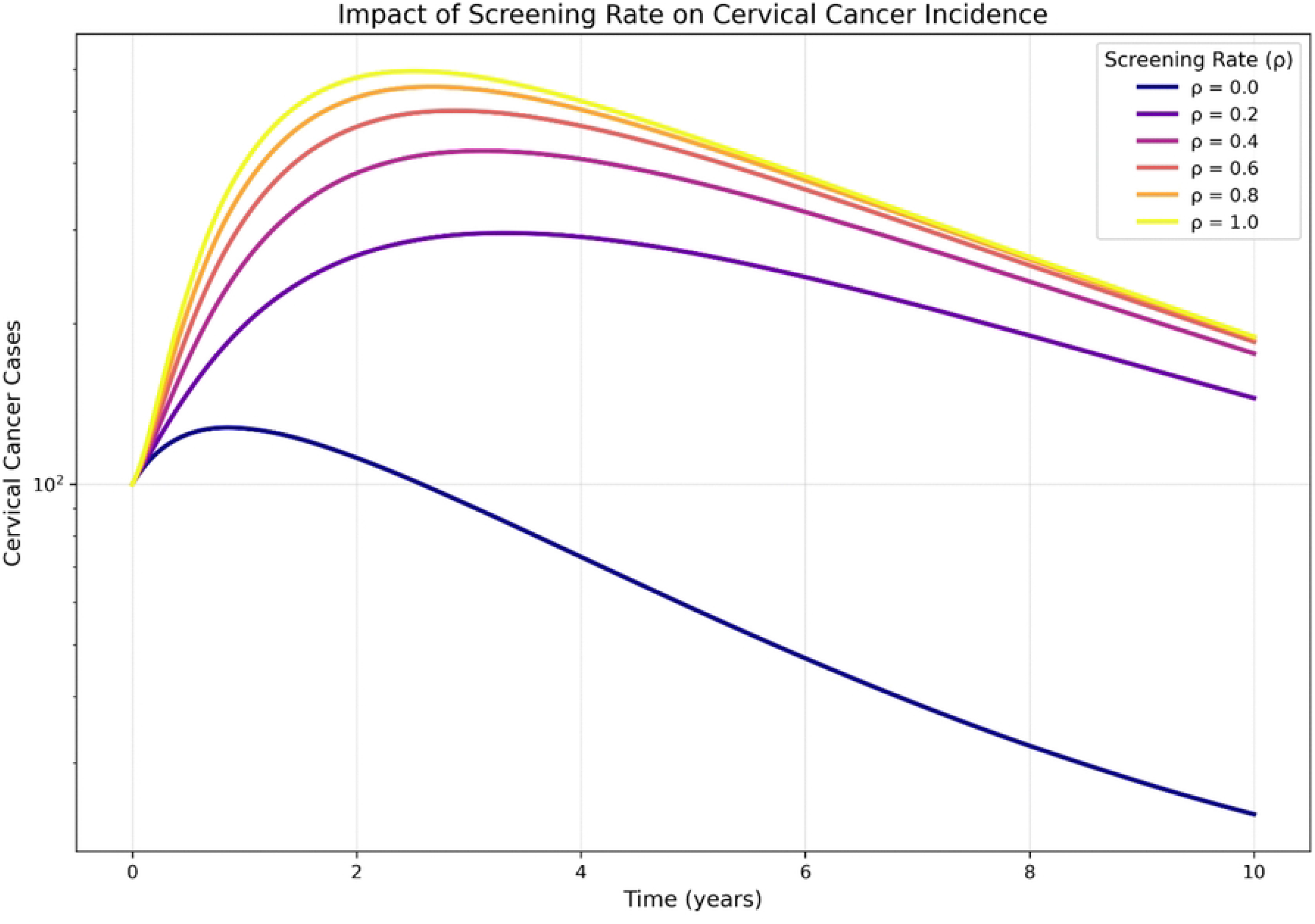
Varying screening rates on Cervical cancer incidence.

Fig 10 and Fig 11 displays how different vaccination coverage levels affect the number of cervical cancer cases ( Fig 10 ) and HPV cases ( Fig 11 ) over time. When *θ* = 0.0, highest Cervical cancer cases and high HPV prevalence throughout the simulation. This represent the natural progression of HPV to Cervical cancer. When *θ* = 1.0, HPV levels are very low approaching elimination, which leads to no development of Cervical cancer. Fig 12 demonstrates that increasing treatment capacity directly reduces Cervical cancer prevalent. High screening rates (*γ*) lowers Cervical cancer incidence while high cancer treatment (*α*) shows better survival rates. High treatment rates alone are less effective if the screening rates is low, as many cases remain undetected until late stages. Fig 13 and Fig 14 demontrates the impact of treatment on Cervical cancer cases and HPV prevalence.

**Fig 10.**
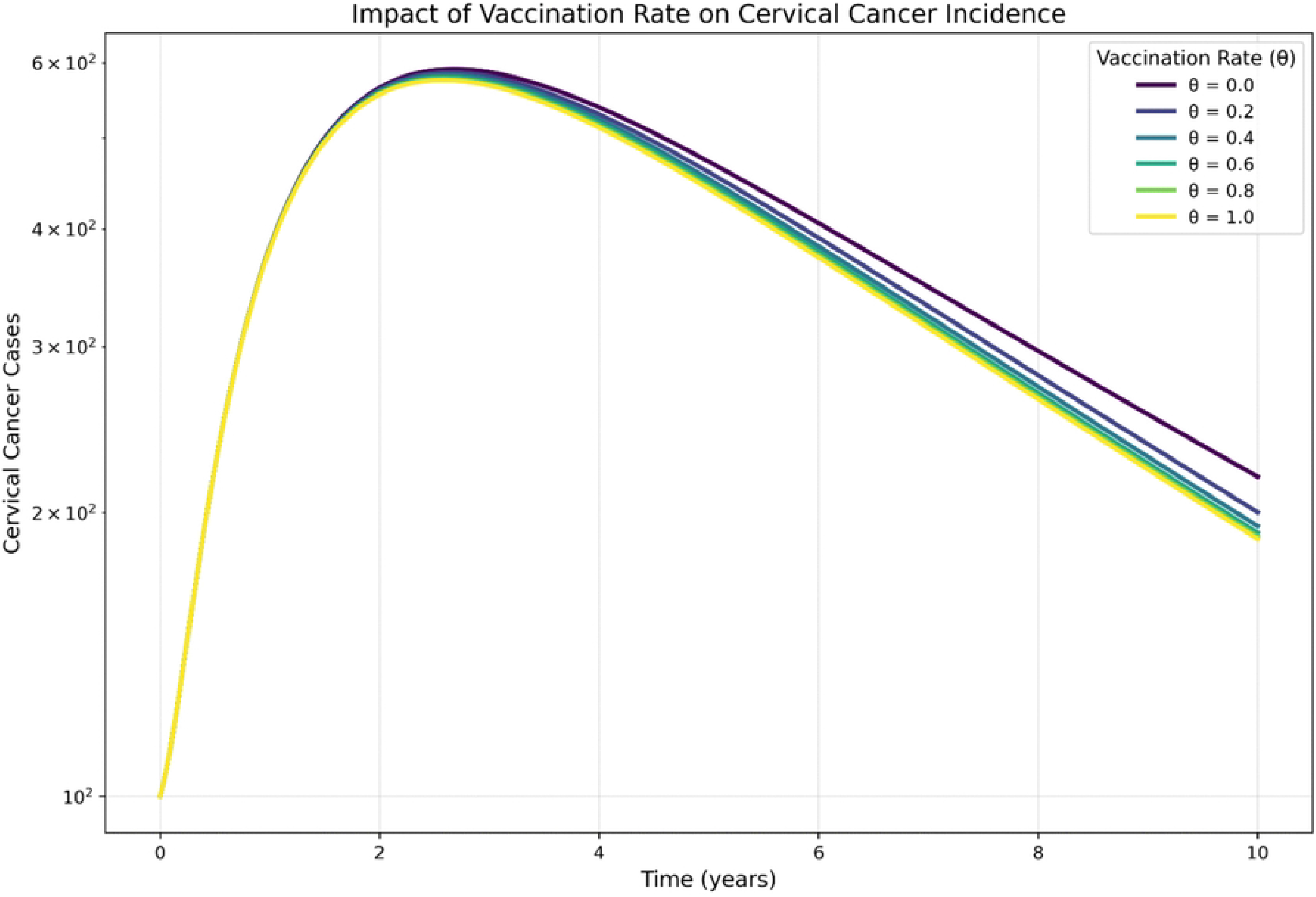
Varying vaccination rates on Cervical cancer cases.

**Fig 11.**
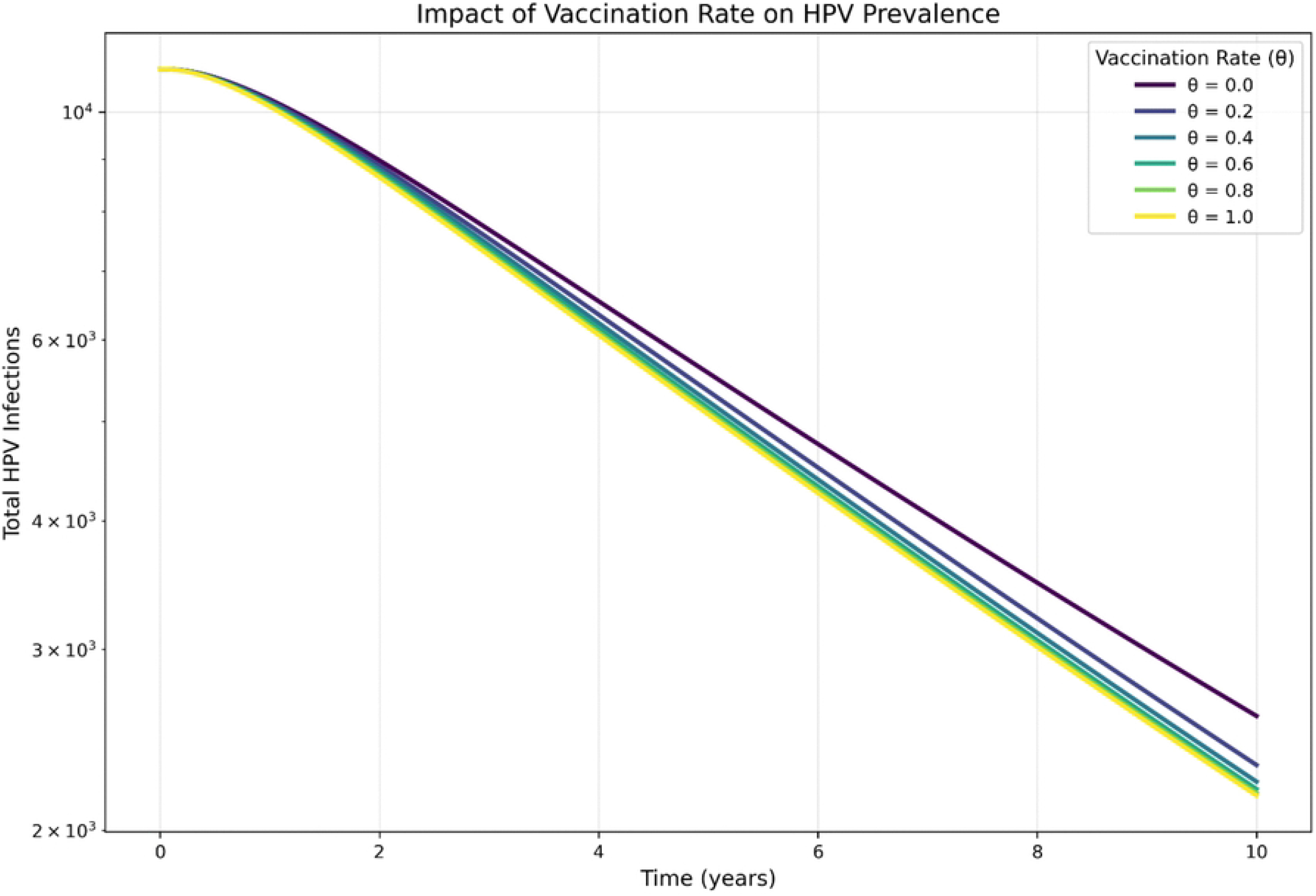
Varying vaccination rates on HPV incidence.

**Fig 12.**
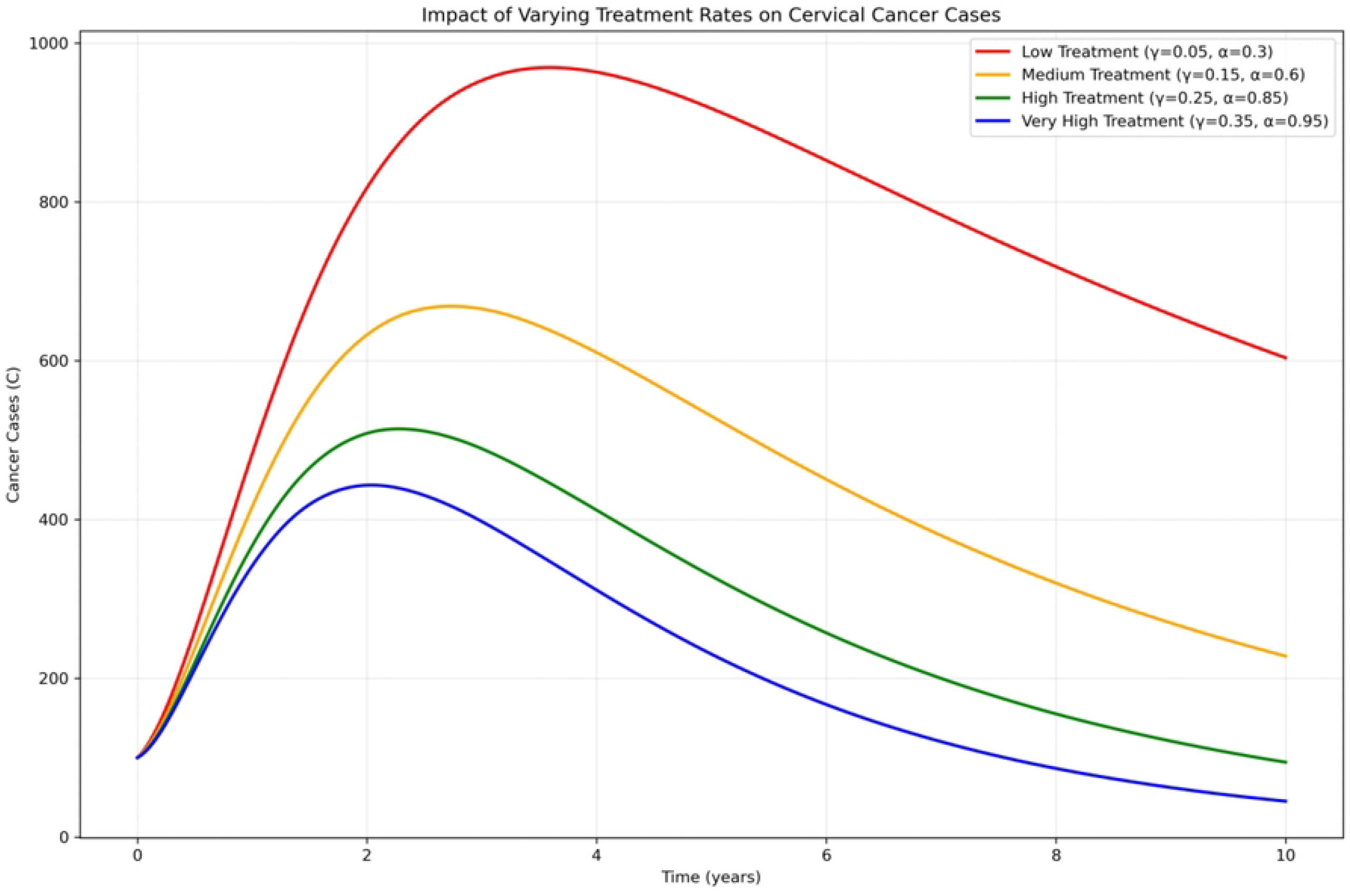
Varying screening and treatment on cervical cancer.

**Fig 13.**
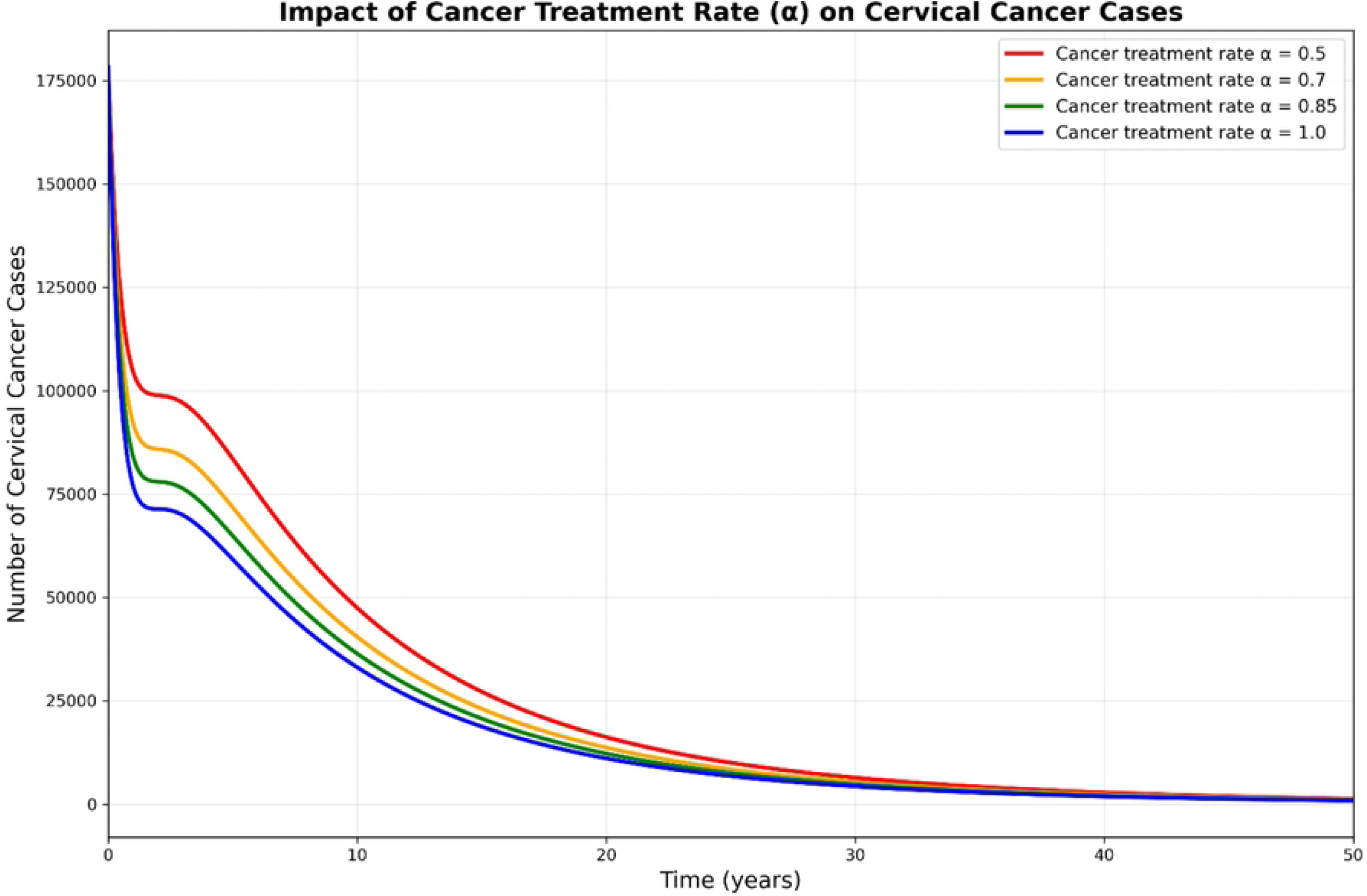
Impact of Treatment on Cervical cancer cases.

**Fig 14.**
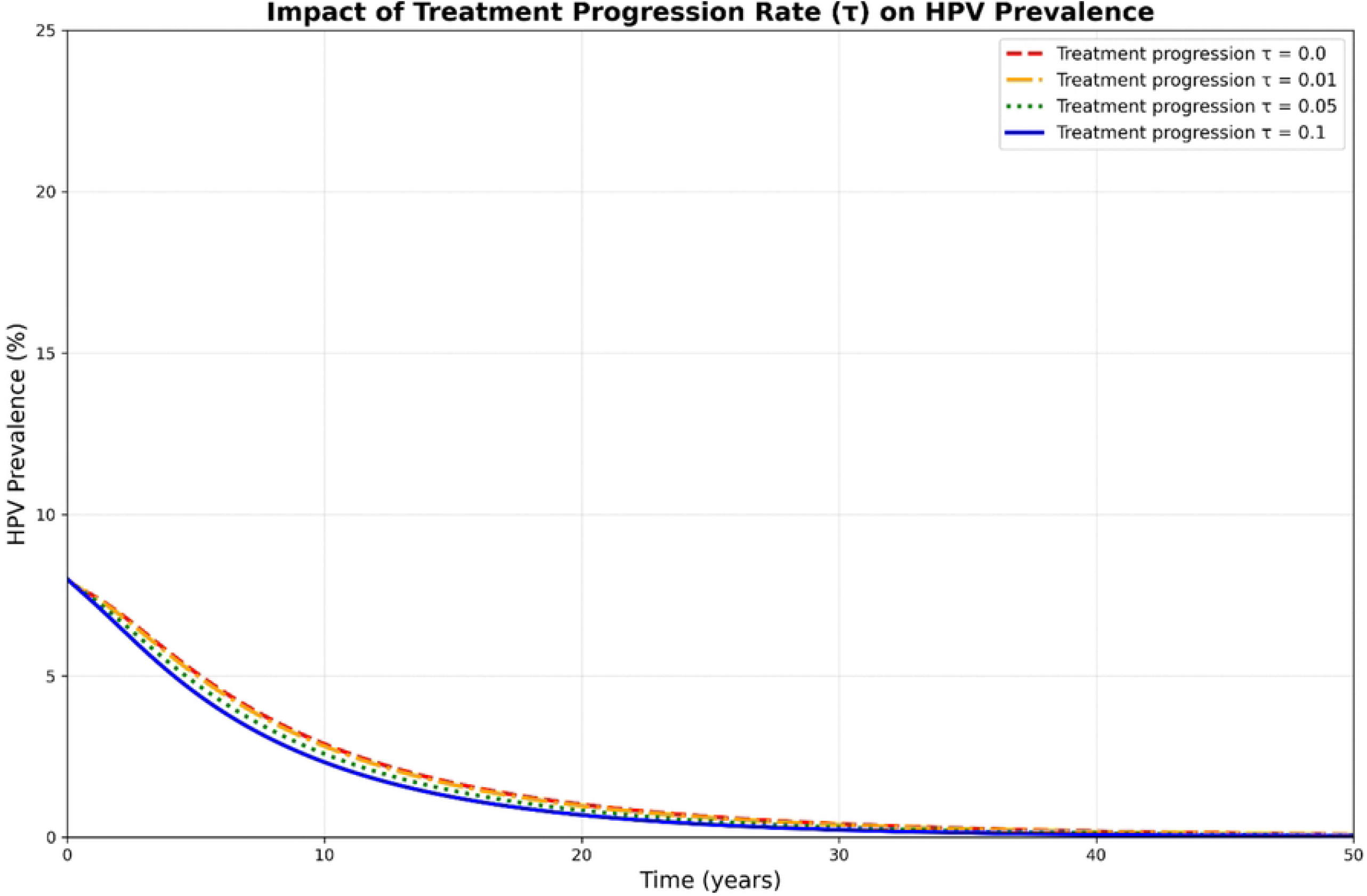
Treatment rates affect HPV prevalence.

### Conclusions

In conclusion, a mathematical model to investigate the impact of screening, vaccination and treatment on the transmission dynamics of HPV-Cervical cancer has been developed. We proved the model solutions positivity and boundedness properties in a biologically feasible region. Additionally, we computed the equilibrium points of the model and analysed their stability with respect to the basic reproduction number. We proved that the disease-free stationary states are stable provided that the reproduction number is less than one. The unique endemic equilibrium is locally asymptotically stable if reproduction number is greater than one and unstable otherwise. The endemic equilibrium points exist, and a backward bifurcation also occurs, indicating that *R*_0*c*_ *<* 1 is a necessary but not sufficient condition for eradicating HPV-Cervical cancer from the community. Moreover, the results from sensitivity analysis show that an increase in vaccination,screening and treatment leads to reduction in the reproduction number . Finally the results from the numerical simulations imply an effectual decrease of the transmission rate and and a simultaneous increase of vaccination,screening and treatment leads to the minimizing the spread of HPV-Cervical cancer, as we had seen in the numerical Fig 5 to Fig 14. Moreover, we want to extend to apply optimal control problem and cost effectiveness to advice optimal strategies for mitigating the disease in the community.

## Data Availability

All relevant data are within the paper

